# Metabolic phenotyping of tear fluid as a prognostic tool for personalised medicine exemplified by T2DM patients

**DOI:** 10.1101/2021.11.30.21267045

**Authors:** Julia Brunmair, Andrea Bileck, Doreen Schmidl, Gerhard Hagn, Samuel M. Meier-Menches, Nikolaus Hommer, Andreas Schlatter, Christopher Gerner, Gerhard Garhöfer

## Abstract

**Background/Aims:** One goal of predictive, preventive, and personalised medicine is to improve the prediction and diagnosis of diseases, as well as to monitor therapeutic efficacy and to tailor individualised treatments with as little side effects as possible. New methodological developments should preferably rely on non-invasively sampled biofluids like sweat and tears in order to provide optimal compliance. Here we have thus investigated the metabolic composition of human tears in comparison to finger sweat and evaluated whether tear analyses may provide insight into ocular and systemic disease mechanisms.

**Methods:** In addition to finger sweat, tear fluid was sampled from 20 healthy volunteers using commercially available Schirmer strips. Tear fluid extraction and analysis using high-resolution mass spectrometry hyphenated with liquid chromatography was performed with optimized methods each for metabolites and eicosanoids. As second approach, we performed a clinical pilot study with 8 diabetic patients and compared them to 19 healthy subjects.

**Results:** Tear fluid was found to be a rich source for metabolic phenotyping. Remarkably, several molecules previously identified by us in sweat were found significantly enriched in tear fluid, including creatine or taurine. Furthermore, other metabolites such as kahweol and various eicosanoids were exclusively detectable in tears, demonstrating the orthogonal power for biofluid analysis in order to gain information on individual health states. The clinical pilot study revealed that many endogenous metabolites that have previously been linked to type 2 diabetes such as carnitine, tyrosine, uric acid and valine were indeed found significantly up-regulated in tears of diabetic patients. Nicotinic acid and taurine were elevated in the diabetic cohort as well and may represent new biomarkers for diabetes specifically identified in tear fluid. Additionally, systemic medications like metformin, bisoprolol, and gabapentin, were readily detectable in tears of patients. These findings highlight the potential diagnostic and prognostic power of tear fluid analyses, in addition to the promising methodological support for pharmacokinetic studies and patient compliance control.

**Conclusions:** Tear fluid analysis may support the development of clinical applications in the context of predictive, preventive, and personalised medicine as it reveals rich molecular information in a non-invasive way.

## Introduction

In clinical metabolomics applications, blood sampling and analyses are typically employed in order to identify biomarkers for diagnosis and prognosis. However, blood collection is invasive and thus, impedes time-course measurements and the identification of dynamic biomarkers due to several compliance issues as multiple samples would have to be collected in short intervals.[1-3] This calls for the evaluation of alternative body fluids, such as sweat, saliva, or tears.[4, 5] All of these matrices are accessible in a non-invasive fashion, and can be collected painlessly, rapidly, with only minimal to no discomfort and stress for the patients, thus, supporting optimal compliance for biomedical studies.[1, 6-9] As we have previously investigated, the composition of sweat collected from the fingertip was found to be a rich source for various biologically highly relevant metabolites.[10, 11] Sweat from fingertips can be sampled in a frequent manner (up to 12 times per hour), hence, finger sweat analysis proved to be an optimal tool for kinetic measurements, exemplified by the ingestion of caffeine and its hepatic conversion to paraxanthine, theobromine and theophylline. However, sweat analysis faces a normalisation problem impeding absolute quantification of metabolites as the exact amount of collected sweat cannot be determined. The implementation of mathematical models of related metabolic pairs measured in sweat allowed to overcome this limitation and permit the calculation of the sweat rate for each sample.[11] Further, sweat and also saliva may suffer from contamination stemming from the skin and bacteria, food intake or smoking, respectively, potentially impeding and distorting highly sensitive mass spectrometry (MS)-based metabolomic analyses. [1, 4] In contrast to saliva and sweat, tear fluid also faces the least contamination problems and tear volume can be determined when tear fluid is collected with the commercially available Schirmer strips. However, tears cannot be sampled as frequently as sweat or saliva as Schirmer strips may irritate the eye. [1, 12] Thus, we decided to evaluate the biomedical power of tear fluid analysis in a more systematic fashion.

### Tear fluid composition and its potential applications in the context of PPPM

Due to its distinct origin, tears may actually contain biomarker candidates hardly accessible via blood or sweat. The function of the ocular tear film is complex including maintenance of lubrication of the ocular surface, guaranteeing normal vision and immune defence of the eye. [7, 13-15] Compared to sweat, which is mainly excreted by eccrine and apocrine glands, tears get secreted from different glands: Whereas the lacrimal glands mainly produce the aqueous component of the tear film, the meibomian glands, goblet cells, and ocular surface epithelial cells are responsible for the lipid and mucin components of the tear film.[7, 14] Next to water (98%)[13], tear fluid comprises proteins such as lipocalin, lactotransferrin, lysozyme C and immunoglobulins [16-19], lipids (e.g. phospholipids, sphingolipids, wax esters and triglycerides), glycans, electrolytes (e.g. sodium, potassium, chloride and phosphate)[20], as well as metabolites like amino acids, urea, cholesterol, creatine, and epinephrine.[21] Changes in tear film composition have already been explored in many ocular diseases such as age-related macular degeneration, dry eye disease, glaucoma or diabetic retinopathy using ‘Omics’ approaches. Additionally, systemic diseases like cancer, multiple sclerosis, Alzheimer’s disease, and diabetes have also been linked to alterations of the molecular composition of tears.[7, 20, 22-31] Several medications such as antibiotics, chemotherapeutics, and anti-inflammatory drugs were further successfully detected in tears after drug administration.[32-36] Regarding tear fluid analysis, metabolomics is still rarely applied compared to other ‘Omics’ techniques. Thus, the metabolic investigation of tear fluid may support the identification of novel biomarkers for diagnosis and prognosis or the monitoring of treatment efficacy, allowing for a more patient-tailored therapy.[37]

### Metabolomic investigations supporting biomarker discovery of type 2 diabetes mellitus

Type 2 diabetes mellitus (T2DM) is a metabolic disorder affecting half a billion of patients worldwide.[38] However, the underlying disease mechanisms and potential risk factors are not yet fully elucidated. Non-modifiable risk factors like genetic predisposition and advanced age, but also modifiable risk factors associated with the personal lifestyle such as physical (in-)activity and sugar-rich diet, leading to elevated Body Mass Index (BMI) and increased fasting plasma glucose levels, are synergistically involved in diabetes manifestation.[38, 39] T2DM is not only associated with premature mortality, but also several other health issues including kidney failure, retinopathy, increased risk of cardiovascular disease and stroke as well as with a reduced quality of life.[40, 41] Predictive biomarkers are urgently needed for early identification of individuals at high risk of developing T2DM in order to design highly-effective targeted preventive measures.[42] Untargeted metabolomics of plasma samples from diabetic patients has already proven to provide novel insights into mechanisms of disease pathogenesis and to supply biomarkers for diabetes.[43] As diabetes is a systemic disease that is known to also affect the eye (diabetic retinopathy), it was presently chosen as model disease for the evaluation of tear fluid in a clinical context.

### Tear fluid analysis enabling the metabolic phenotyping of T2DM

In this basic research project, we have developed a simple and quick sample preparation workflow for the comprehensive investigation of tear fluid, which is based on our previously established liquid chromatography-mass spectrometry (LC-MS) methods for sweat metabolomics [10, 11] and plasma eicosadomics. [44, 45] First, we wanted to systematically investigate and compare tear fluid composition with sweat in order to find differences deriving from the respective secretory glands. Indeed, distinct small molecules were preferably accumulated in tears such as eicosanoids, taurine or epinephrine, while other molecules like amino acids could be found in higher concentrations in sweat, demonstrating that each fluid has its unique set of metabolites. These findings prove tear fluid as a rich metabolic source being worth for further investigations regarding biomarker discovery. Therefore, as a second part of this study, the potential of tear fluid analysis for biomarker discovery in T2DM was investigated. To this end, exogenous as well as endogenous metabolites of diabetic patients and healthy controls were profiled in tear fluid. Remarkably, many blood-borne metabolites that have already been linked to diabetes such as tyrosine, valine, carnitine, and uric acid were presently found significantly upregulated in the tears of diabetic patients. Moreover, we have found individual medication (*e.g*. metformin), but also lifestyle factors (*e.g*. the sweetener sorbitol) associated with the disease in the tear fluid of diabetic patients. Thus, here we demonstrate the potential of tear fluid analyses for diagnosis but also for lifestyle monitoring, potentially supporting the development of novel applications in the context of predictive, preventive and personalised medicine (PPPM).

## Materials and Methods

### Reagents and Chemicals

LC-MS grade formic acid, water, and acetonitrile used for chromatographic separation as well as for preparation of internal standards and samples were purchased from VWR (Germany), whereas LC-MS grade methanol was bought from Honeywell International Inc. (USA). Internal and external standards used for normalisation and verification were obtained either from Sanova Pharma GmbH (Austria) or Sigma Aldrich (Austria). Filter papers used for the collection of sweat from fingertips were stamped out of fuzz free paper (precision wipes, number=7552, white, 11×21cm, Kimtech Science, Kimberly-Clark Professional, USA). Tears were collected using Schirmer plus paper strips (Grecis, France).

### Cohort Design

Volunteers were recruited by Department of Clinical Pharmacology, Medical University of Vienna, Austria, and gave their written, informed consent for the different studies outlined in Table 1. All experiments were approved by the ethics committee of the Medical University Vienna. For study A sweat from the fingertips and tear fluid from both eyes were sampled in parallel, whereas for study B and C only tears were obtained. Some volunteers were sampled several times. In case of the diabetic participants, basic clinical and ophthalmological characteristics were collected (Table 2). To diagnose whether patients suffered from diabetic retinopathy, pupils were dilated with tropicamide 0.5% eye drops for better visualisation during eye examination. For the assessment of intraocular pressure, a mixture of oxybuprocaine and fluorescein was used. Moreover, patients were asked about medication they took on a regular basis, of which some could be detected in tears.

**Table 1:** Overview of all studies discussed in this publication. Age and BMI are expressed as average ± standard deviation.

| Study | Participants | Age | BMI [kg m<sup>-2</sup>] | Sampling |
| --- | --- | --- | --- | --- |
| **A** | 20 healthy subjects | 31 ± 13 | 25 ± 4 | Sweat and tears, two visits |
| **B** | 20 healthy subjects | 24 ± 6 | 22 ± 6 | Tears, up to 5 visits |
| **C** | 8 diabetic patients | 64 ± 9 | 32 ± 4 | Tears, 1 visit |

**Table 2:**
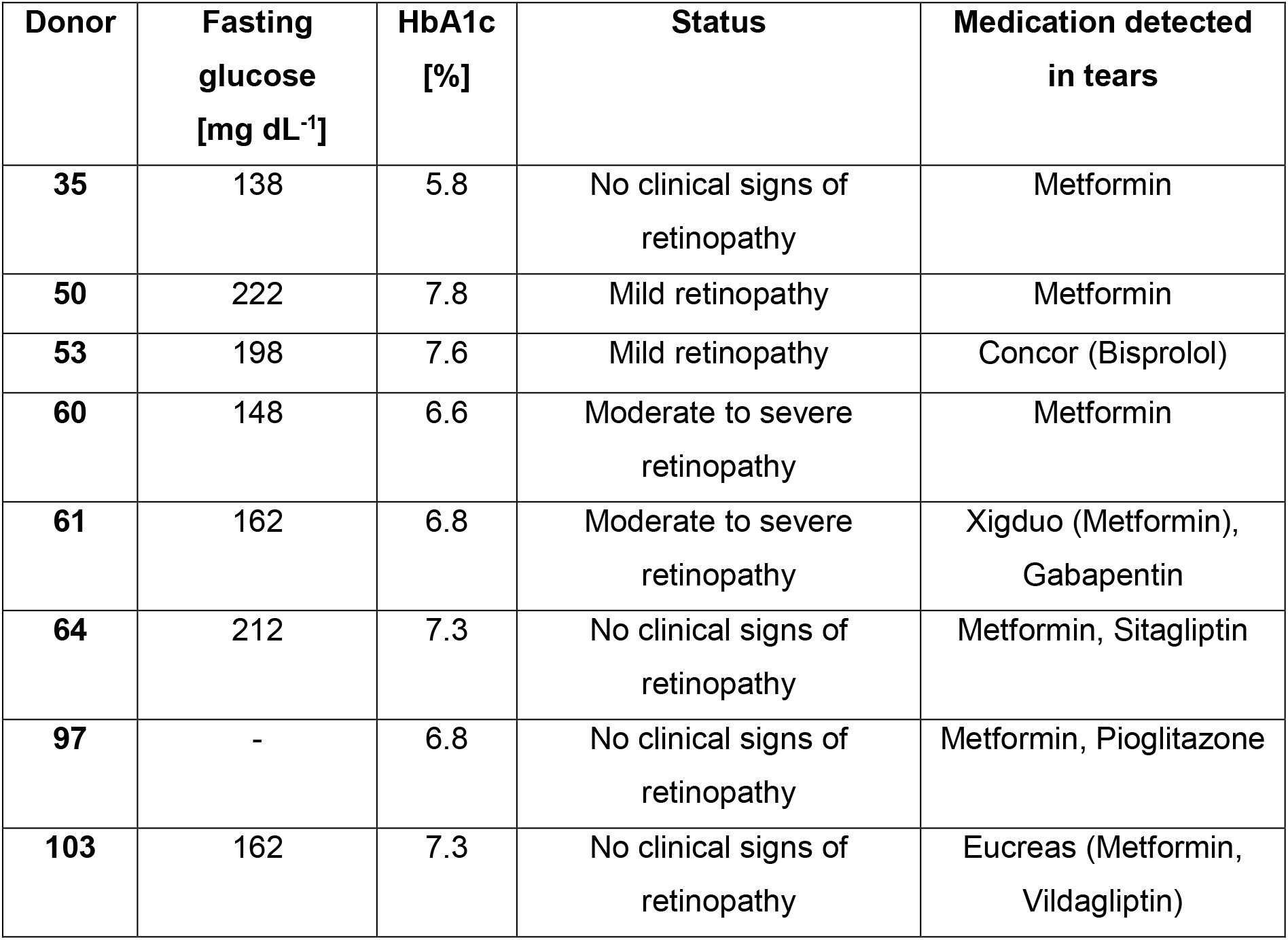
Clinical characteristics for diabetic patients

| Donor | Fasting glucose [mg dL<sup>-1</sup>] | HbA1c [%] | Status | Medication detected in tears |
| --- | --- | --- | --- | --- |
| **35** | 138 | 5.8 | No clinical signs of retinopathy | Metformin |
| **50** | 222 | 7.8 | Mild retinopathy | Metformin |
| **53** | 198 | 7.6 | Mild retinopathy | Concor (Bisprolol) |
| **60** | 148 | 6.6 | Moderate to severe retinopathy | Metformin |
| **61** | 162 | 6.8 | Moderate to severe retinopathy | Xigduo (Metformin), Gabapentin |
| **64** | 212 | 7.3 | No clinical signs of retinopathy | Metformin, Sitagliptin |
| **97** | - | 6.8 | No clinical signs of retinopathy | Metformin, Pioglitazone |
| **103** | 162 | 7.3 | No clinical signs of retinopathy | Eucreas (Metformin, Vildagliptin) |

### Collection of Sweat from the Fingertip and Sample Preparation

Finger sweat samples were collected and processed as previously described.[10, 11] In short, pre-wetted filter papers were provided in 0.5 mL Eppendorf tubes. For each sweat collection, volunteers washed their hands with warm tap water and dried them with disposable paper towels. After waiting for 1 min at room temperature without touching anything, filter papers were placed between the thumb and index finger using clean tweezers and held for another minute, resulting in the collection of 2 min sweat in total. Then, the sampling unit was transferred into the labelled Eppendorf tube and stored at 4° C. For the extraction of the metabolites, an extraction solution consisting of an aqueous solution of caffeine-(trimethyl-D9) in a concentration of 1 pg µL^-1^ with 0.2% formic acid was prepared. 120 µl of this extraction solution were added into the 0.5 mL Eppendorf tubes containing the sampling unit. The metabolites were extracted by pipetting up and down 15 times. The filter paper was pelleted on the bottom of the tube and the supernatant was transferred into HPLC vials equipped with a 200 µL V-shape glass insert (both Macherey-Nagel GmbH & Co.KG) and analysed by LC-MS/MS.

### Collection of Tear Fluid and Sample Preparation

Tear fluid was collected according to the instructions of the manufacturer. Briefly, donors were asked to look up, then the lower eyelid was gently pulled down and the bent strip end of the Schirmer plus strip was placed in the temporal section of the lower conjunctival fornix. Both eyes were tested at the same time. The donors were then asked to gently close their eyes without squeezing. After a maximal collection time of 5 minutes, subjects were instructed to open their eyes again, and the strips were removed. Schirmer strips were transferred into 2 ml glass vials (Agilent Technologies, USA) after tear collection and stored at -20 °C until further sample preparation. Different extraction solution and reconstitution conditions were evaluated to determine the best conditions for the parallel analysis of eicosanoids and metabolites: i) 200 µL water with 0.2% formic acid and internal standards without pre-concentration and reconstitution, ii) 300 µL 35% methanol with 0.2% formic acid and internal standards without pre-concentration and reconstitution, iii) 300 µL 80% methanol and internal standards followed by drying and reconstitution in 5% methanol (s. results). Finally, samples were prepared as follows: Filter papers were first thawed before 5 µL of a deuterated eicosanoid standard mixture containing 15S-HETE-d8, 12S-HETE-d8, 5-oxo-ETE-d7, PGE2-d4, 20-HETE-d6, and 11,12-DiHETre-d11 (Cayman Europe, Tallinn, Estonia) were pipetted onto Schirmer strips. Exact concentrations for the deuterated eicosanoid standards can be found in Supplementary Table 1. For the extraction of the samples, 300 µL of 80% methanol were added onto the filter paper, samples were vortexed and additionally, put on a shaker for 30 min. The Schirmer strip was pelleted at the bottom of the glass tube and the supernatant was transferred into a new HPLC glass vial (Macherey-Nagel GmbH & Co.KG). Samples were dried using a gentle stream of nitrogen and reconstituted in 200 µL of 5% methanol, 0.2% formic acid containing the internal standards caffeine-(trimethyl-D9) and N-acetyl-tryptophan (both in a concentration of 1 pg µL^-1^). Samples were transferred into V-shaped glass inlets, put back in the labelled HPLC glass vial and analysed by LC-MS/MS. Schirmer strips with no tears were subjected to the same extraction protocol to serve as blank controls.

### Untargeted LC-MS/MS Analysis

For LC-MS/MS analysis the Q Exactive HF mass spectrometer coupled to a Vanquish UPLC system (both Thermo Fisher Scientific) was used and both were controlled by the Xcalibur software (Thermo Fisher Scientific).

For the metabolomics analyses of both sweat and tears, chromatographic separation was achieved using a Kinetex XB-C18 column (100 Å, 2.6 µm, 100 × 2.1 mm, Phenomenex Inc.). Mobile phase A was water with 0.2% formic acid and mobile phase B was methanol with 0.2% formic acid. The following gradient was used: 0.0-0.3 min 1-5% B, then 0.3 – 4.5 min 5-40% B, followed by a column washing phase from 4.5 – 6.9 min at 80% B and then a re-equilibration phase of 1.6 min at 1% B resulting in a total runtime of 7.5 min. The column temperature was set to 40°C, the flow rate was 500 µL min^-1^, and the injection volume was 10 µL. Samples were analysed in technical duplicates. An untargeted mass spectrometric approach was applied for compound identification. Therefore, electrospray ionisation was performed in the positive mode, the MS scan range was from 100-1000 *m/z* and the resolution was set to 60000 (at *m/z* 200). A top 4 method was applied and dynamic exclusion was set to 6 s. Selected precursors were fragmented applying 30 eV collision energy and fragments were subsequently analysed in the orbitrap at a resolution of 15000 (at *m/z* 200).

For eicosanoid profiling of sweat and tears, a Kinetex C18 column (2.6 μm, 100 Å, 150 × 2.1 mm; Phenomenex Inc.) was used, and a 20 min gradient flow method was applied to separate molecules. Mobile phase A was again water with 0.2% formic acid, whereas mobile phase B consisted of 90% acetonitrile, 10% methanol, and 0.2% formic acid. The gradient was programmed as follows: starting with 35% B for 1 min, then 35-90% B from 1-10 min, followed by a wash phase at 99% B for 3.25 min before returning to the starting condition of 35% B. The column temperature was set to 40°C, the flow rate was kept at 200 µL min^-1^, and the injection volume was 20 µL. Eicosanoids were measured in negative ionisation mode and the scan range for MS1 spectra was from 250-700 *m/z* at a resolution of 60000 (at *m/z* of 200). The two most abundant precursor ions were selected for fragmentation, applying a collision energy of 24 eV. An inclusion list (Supplementary Table 2) for prioritized fragmentation was predefined for *m/z* corresponding to well-known eicosanoids and their precursors. Generated fragment ions were analysed in the orbitrap at a resolution of 15000 (at 200 *m/z*).

### Data Analysis, Statistics and Figures

Raw files generated by the Q Exactive HF instrument were processed by the Compound Discoverer Software 3.1 (Thermo Fisher Scientific) using a user-defined workflow tree. Automatically identified compounds were manually reviewed using Xcalibur 4.0 Qual browser (Thermo Fisher Scientific) and additionally, in case of metabolites the obtained MS2 spectra were compared to a spectral library (mzcloud - Copyright © 2013–2021 HighChem LLC, Slovakia). For eicosanoid identification, recorded MS2 spectra of eicosanoid specific precursor masses were compared to reference spectra available in the LipidMaps spectral library and mzcloud. The match factor cut-off from mzcloud in the Compound Discoverer Software was set to ≥ 80 for manual investigation, and the maximum mass tolerance was 5 and 10 ppm on MS1 and MS2 level, respectively. Verification of key metabolites was done with purchased analytical standards analysed under the same LC-MS conditions as samples. The Tracefinder Software 4.1 (Thermo Fisher Scientific) was used for peak integration and calculation of peak areas. Peaks with a signal-to-noise of >25 and a minimum area of 1E5 were considered for further evaluation. Afterwards, batch tables generated by the Tracefinder Software were exported and further processed by means of Microsoft Excel, GraphPad Prism (Version 6.07) for univariate analysis such as *t*-tests or Mann Whitney tests as well as to check if data display gaussian distribution, and the Perseus software (version 1.6.12.0)[46] for multivariate analysis namely PC and volcano plots. Figure 1A was created using BioRender (www.BioRender.com).

**Figure 1.**
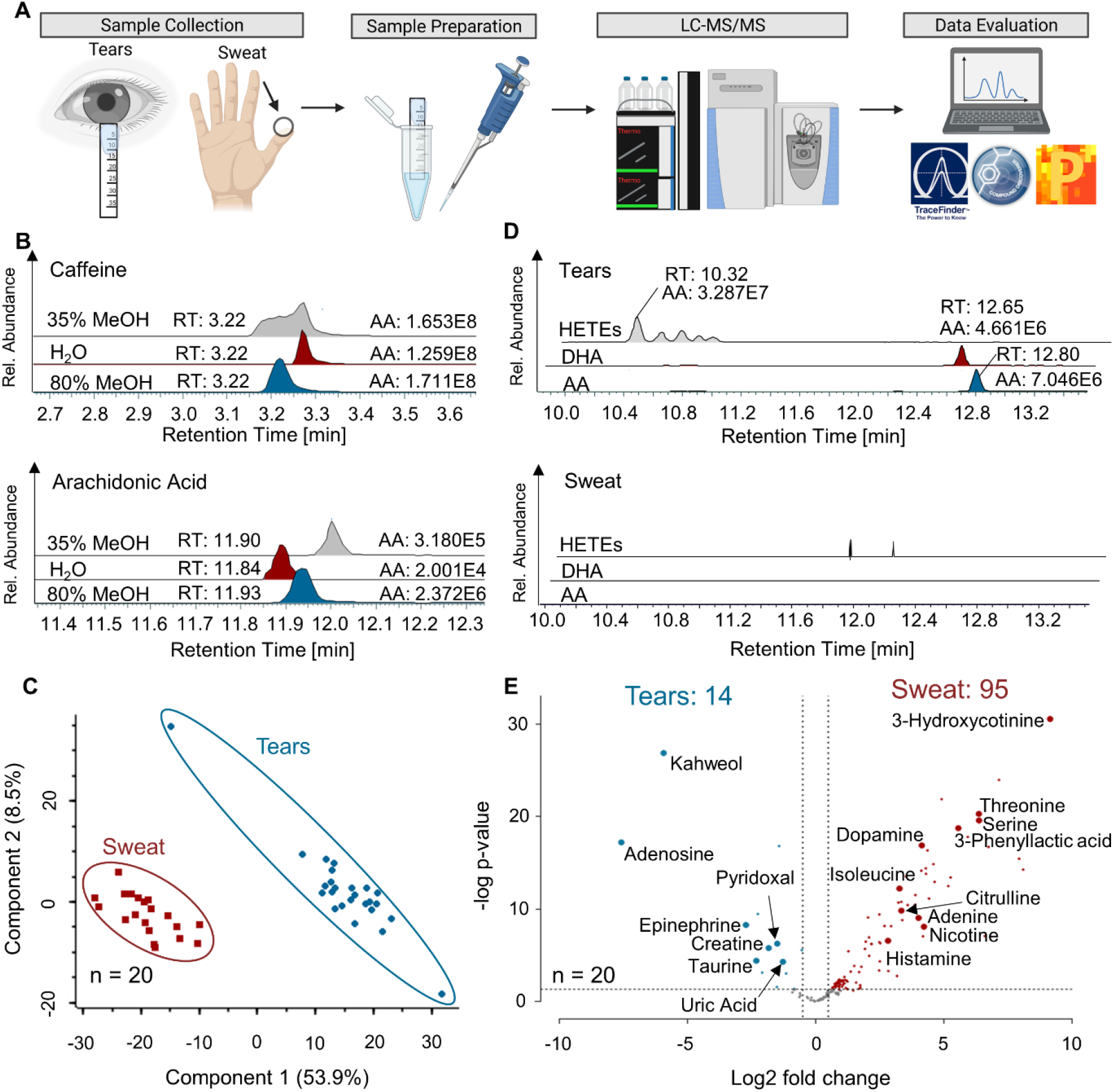
Non-invasively sampled tear fluid and sweat have distinct metabolic compositions. A straight-forward workflow for tear fluid collected with commercially available Schirmer strips was established and successfully applied to proof-of-principle studies. **A** Graphical summary of the workflow including sample collection of tear fluid and sweat from fingertips, the extraction of analytes and subsequent LC-MS/MS analysis as well as data analysis. **B** Respective peak areas of caffeine and arachidonic acid extracted from tear fluid with water, 35% methanol and 80% methanol. AA, absolute area; RT, retention time **C** Principal component analysis (PCA) of metabolites detected in sweat and tears simultaneously derived from the same healthy volunteers is depicted. The PCA was calculated with a set of 165 metabolites identified in both sweat and tears and successfully clustered samples according to biofluids. **D** Examples of eicosanoids (hydroxyeicosatetraenoic acids (HETEs), docosahexaenoic acid (DHA) and arachidonic acid (AA)) identified in high levels in the tear sample of a healthy control (relative intensities of 4E6 – 3E7), but not in the respective sweat sample. AA, absolute area; RT, retention time **E** Metabolic differences of sweat and tears depicted with a volcano plot. 14 and 95 metabolites were found at higher levels or specifically in tears and sweat, respectively.

## Results

### Tear fluid has a distinct metabolic composition compared to sweat

A straight-forward workflow was established for processing tear fluid collected with Schirmer strips according to the manufacturer’s protocol (Figure 1A). In short, metabolites were extracted from Schirmer strips using organic conditions, the resulting solution was evaporated, reconstituted in the initial solvent conditions of the LC method and subsequently analysed by high-resolution MS. Sample collection takes ≤ 5 min depending on tear flow rate of the individual and sample preparation of a single tear fluid can be performed within 5 min. In addition of being fast and simple, sampling of tear fluid is non-invasive, which facilitates patient compliance as it is less discomforting compared to blood collection. LC-MS data acquisition of samples requires further 7.5 min in case of metabolites or 20 min in case of eicosanoids, which gives a total of approximately 40 minutes for the entire workflow per sample.

Metabolic profiling of minute amounts of sweat as well as of low abundant plasma eicosanoids using the Q Exactive HF hyphenated with an ultra-high-performance LC (UPLC) was already successfully demonstrated by us [10, 11, 44, 45], consequently, this set-up was also applied for tear fluid analysis. Regarding sample preparation, several extraction conditions were tested in order to ensure optimal extraction for both, metabolites and eicosanoids, present in tears (Figure 1B). First, water was evaluated as solvent as it is the standard method for sweat analysis. Further, extraction using 35% methanol representing the loading conditions of the eicosanoid analysis method was investigated. Lastly, we evaluated the extraction with 80% methanol followed by drying and reconstitution in 5% methanol, which was reported in the literature.[21] The evaluation of different extraction solutions was performed regarding peak areas and shapes of representative molecules. For caffeine and arachidonic acid (Figure 1B) as well as for paraxanthine and docosahexaenoic acid (Supplementary Figure 1), highest peak areas and best peak shapes were obtained using 80% methanol as extraction solution. Additionally, 80% methanol-extracted preliminary tear fluid led to more feature annotations by the Compound Discoverer Software in contrast to the other two extraction conditions, however, Schirmer strip background levels were similar for all conditions. Thus, 80% methanol was used for sample preparation in the following proof-of-principle studies.

Initially, tear fluid and sweat were collected from 20 healthy donors in order to evaluate and compare the metabolic composition of these biofluids. Many metabolites were detected in both, sweat and tear fluid, but interestingly, a principal component analysis (PCA) using these metabolites (165 in total) separated the two groups perfectly (Figure 1C). Further, tear fluid was found to be a rich source for eicosanoids exemplified by the hydroxyeicosatetraenoic acids (HETEs) and the precursor molecules arachidonic acid (AA) and docosahexaenoic acid. Actually, none of these molecules was detectable in sweat (Figure 1D). Statistical analysis of the metabolites reproducibly detected in all 20 volunteers revealed that 14 metabolites such as creatine, adenosine, taurine, epinephrine and uric acid were found at significant higher levels in tears, whereas other 95 metabolites like adenine, dopamine, nicotine, and histamine were detected at higher levels in sweat (Figure 1E). In case of taurine, abundance levels detected in tear fluid were also significantly higher than in plasma.[24] Interestingly, some metabolites could only be specifically measured in one of the investigated biofluids such as 3-hydroxycotinine and 3-phenyllactic acid in sweat, whereas kahweol and eicosanoids could only be detected in tear fluid (Figure 1D and 1E). These findings indicate that the simultaneous collection and analyses of several biofluids may be synergistic, as some molecules may preferably accumulate in one biofluid due to distinct environmental factors such as pH (sweat: 4.0 – 6.0, tears 7.0 – 8.0 and blood: 7.35-7.45), or polarity. Thus, certain diseases may give rise to biomarkers that are transported to specific biofluids or secreted *via* specific glands.[1] The monitoring of biofluid-specific metabolic responses to certain environmental conditions may consequently gain great relevance in the further development of PPPM.

### Tear fluid analysis allows to discriminate between diabetic patients and healthy controls

In a clinical pilot study, 20 healthy volunteers and 8 T2DM patients were recruited to evaluate tears as a diagnostic and prognostic fluid (Figure 2A). All of these 28 donors received eye drops in the course of their ophthalmological examination, namely oxybuprocaine/fluorescein (Thilorobin ®) for measurement of intraocular pressure (Both study B and C) and tropicamide (Mydriaticum ‘Agepha’ 0.5%) for pupil dilation in diabetes patients (study C). Additionally, tear fluid from both eyes was sampled using Schirmer strips, which were extracted and subsequently analysed by LC-MS/MS. Some of the healthy volunteers were sampled several times, thus, leading to the collection of 127 healthy and 16 diabetic volunteer profiles. Our untargeted metabolomic and eicosadomic workflow applied to the 143 Schirmer strip samples resulted in the identification of 226 metabolites and 70 eicosanoids and eicosanoid-like features (Supplementary Data) in tears of study participants (Figure 2B), including oxybuprocaine, which was administered to all subjects for diagnostic purposes (Figure 2C).

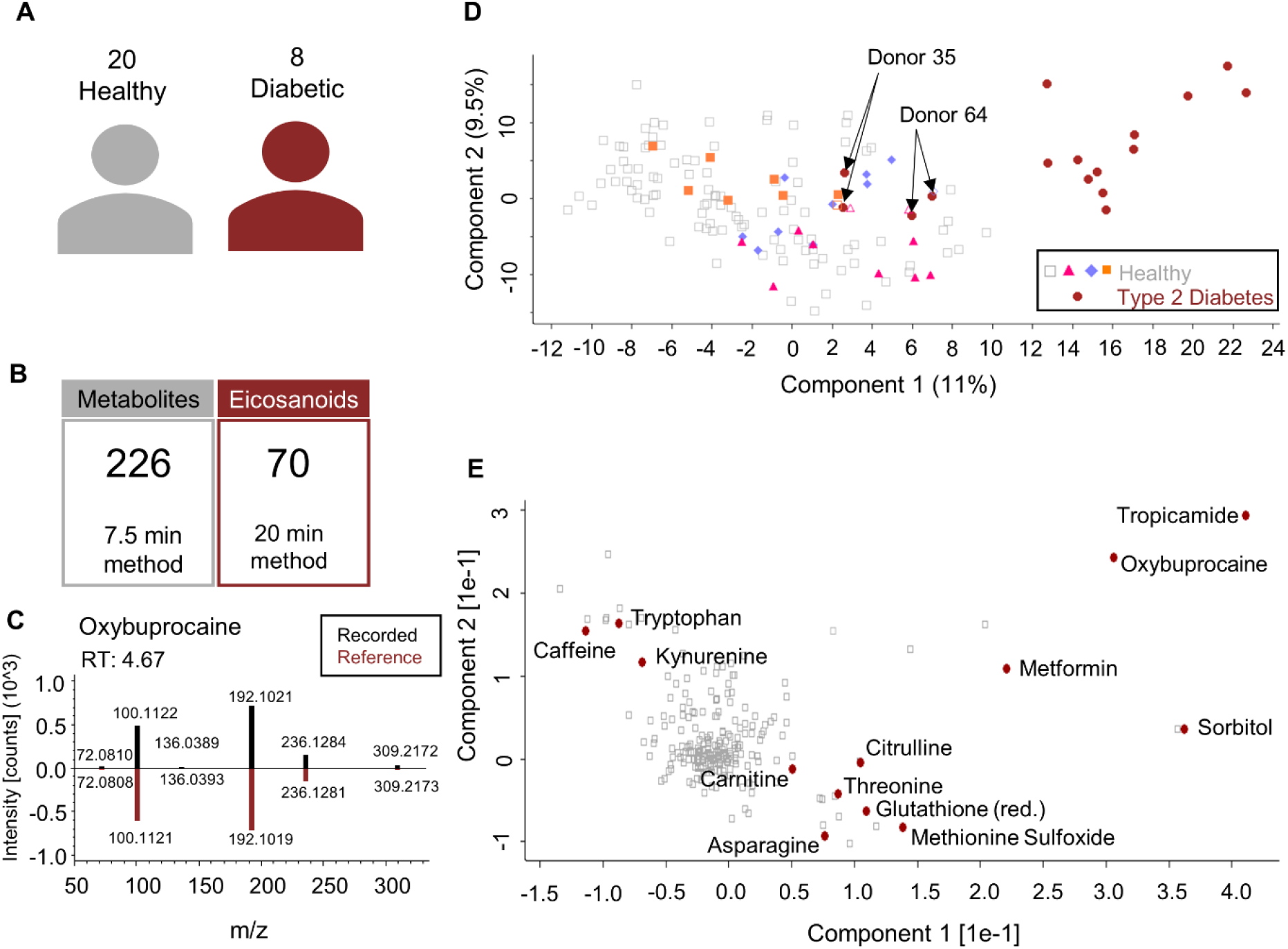

three healthy donors that donated tears more than once (violet, pink and orange) are highlighted.

A PCA using the 226 metabolites successfully discriminated between healthy controls and diabetic patients, however, donors 35 and 64, both belonging to the diabetic cohort, rather clustered with healthy controls (Figure 2D). Interestingly, these two donors showed no signs of diabetic retinopathy according to the clinical evaluation, which may be a causal factor influencing the PCA separation. In addition, some healthy participants donated tear samples at different site visits. Multiple sample of three donors are highlighted in the PCA (Figure 2D, orange, violet and pink symbols), demonstrating that tear samples cluster according to each individual and not the sampling time point. This indicated that the molecular composition of tears collected from both, left and right eye, was determined by the individuum showing little variance associated with multiple sampling. The principal components underlying the PCA plot were influenced by specific medications such as metformin and tropicamide, which was used for pupil dilation during eye examination, but also endogenous factors like methionine sulfoxide, glutathione, threonine and citrulline (Figure 2E). Even though high levels of eicosanoids have been measured in tears (Figure 1), a PCA based on the 70 identified eicosanoids and eicosanoid-like features in tears was not able to discriminate between healthy controls and diabetic patients (Supplementary Figure 2). No significant eicosanoid differences between the two groups were observed. The successful clustering of healthy controls *versus* diabetic patients (Figure 2D) already demonstrated the great potential of tear fluid analyses as a tool for PPPM. In particular, the analysis of tear fluid may not only support the discrimination of distinct disease states but also provide a rich source of biologically relevant metabolites potentially serving as future biomarkers.

### Evaluation of tear fluid as potential source for biomarker candidates

The observation that individuals, who repeatedly donated tears, formed clusters in the PCA (Figure 2D) indicated that the metabolic composition of tear fluid is specific for each individual and hardly affected by sample collection and preparation. This motivated us to investigate the differences between the study groups in greater detail in order to identify potential T2DM-related marker molecules in tear fluid. Statistical analysis of the metabolites detected in tears of study participants revealed significantly higher levels of carnitine, nicotinic acid and sorbitol, as well as tropicamide and metformin in diabetic patients (Figure 3A, 3B and 3C). While carnitine and nicotinic acid are two endogenous metabolites, tropicamide as well as metformin represent individual medications and the sweetener sorbitol may represent a life style related xenobiotic compound. Elevated levels of circulating carnitine have already been observed in obesity and insulin resistance and may be an early predictor of developing T2DM [47, 48]. Altered levels of nicotinic acid have not been reported in the literature before, thus, nicotinic acid might represent a potential novel biomarker associated with diabetes.

**Figure 3.**
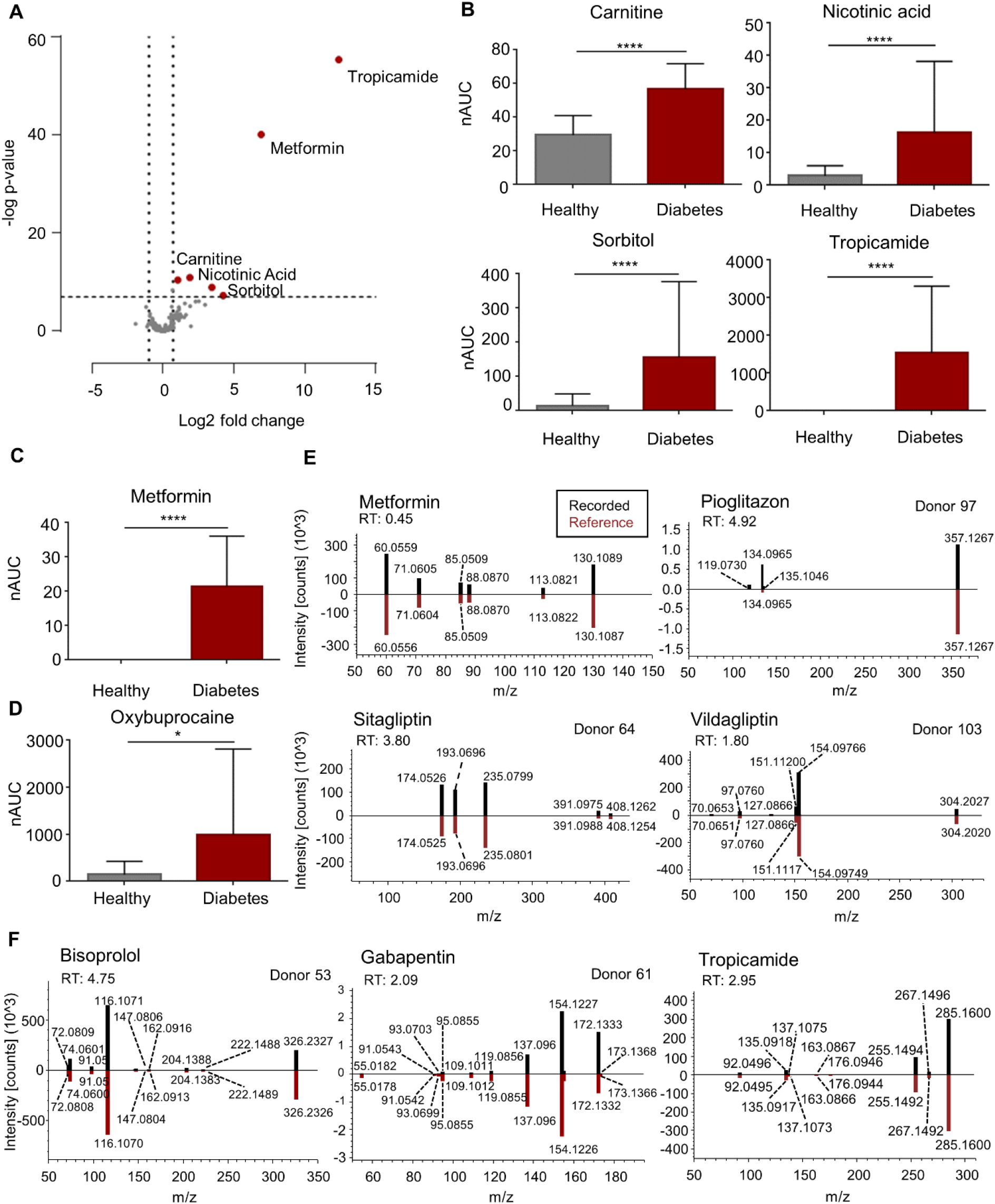
Endogenous and exogenous metabolic differences between diabetic patients and healthy controls. **A** Multiparameter corrected volcano plot revealing significant differences of metabolites between diabetic patients and healthy controls. **B-D** Box plots of carnitine, nicotinic acid, sorbitol, tropicamide (n=127 for healthy and n=16 for diabetes), metformin (n=16) and oxybuprocaine (n=44 for healthy and n=16 for diabetes) are shown for healthy controls and diabetic patients. Since data was not normally distributed, two-tailed unpaired *t*-tests (Mann Whitney test) were performed, demonstrating the significant increase. Oxybuprocaine is only demonstrated for all study participants receiving the same product of eye drops (Thilorbin®, for healthy n = 44, for diabetes n = 16), resulting in a *p*-value of 0.0105. Metformin and tropicamide were only measured in diabetic patients (n = 14), thus, a one sample, two-tailed *t*-test was performed. Boxes are represented as mean and standard deviation. *** = *p*-value of < 0.0001; * = *p*-value of < 0.05; nAUC normalised area under the curve. **E-F** Head-to-tail comparison of recorded *versus* reference spectra taken from *mzcloud* for type 2 diabetes mellitus specific medication metformin (RT = 0.45 min, identified in all diabetic patients except for donor 53), pioglitazone (RT = 4.92 min, only detected in donor 97), sitagliptin (RT = 3.80, only measured in donor 64) and vildagliptin (RT = 1.80, only found in donor 103) as well as for the drugs bisoprolol (RT = 4.75 min, only taken by donor 61), gabapentin (RT = 2.09 min, only consumed by donor 53) and tropicamide (RT = 2.95 min, identified in all diabetic patients). All of them have been identified by the Compound Discoverer Software with a match factor >95 and subsequently verified *via* the patients’ questionnaire. RT, retention time.

Further, a significant difference of oxybuprocaine, the active compound of the eye drops (Thilorobin®), was observed between diabetic patients and healthy (Figure 3D). Oxybuprocaine was found at higher levels in tears of diabetic patients. This observation may indicate that the active ingredient of the eye drops was not as efficiently cleared within the eye in case of diabetic patients compared to healthy individuals. Metabolomic tear fluid analysis actually allowed the identification of individual medications. Next to metformin, which is the first-line oral medication in the treatment of T2DM [49], other diabetic medications such as pioglitazone, sitagliptin or vildagliptin were successfully detected in some patients (Figure 3E). Untargeted mass spectrometric analyses further led to the detection of other drugs in tears of diabetic patients such as the beta-blocker bisoprolol and the anticonvulsant gabapentin, which have indeed been taken by respective patients according to their questionnaire (Figure 3F). Tropicamide, a medication used in diabetic patients during clinical examination in order to assess signs of diabetic retinopathy and to identify potential occurred damage to the retina, was specifically found in the tear fluid of diabetic patients.

These findings clearly demonstrate that tear fluid analyses revealed potential biomarker candidates for diabetes, which have to be further evaluated in larger patient cohorts. Moreover, we have not only identified endogenous molecules, but also xenobiotics that are associated with the disease (e.g. medication and artificial sweetener), allowing to monitor a patient’s compliance regarding the intake of specific prescribed medications.

### Several amino acids and other endogenous molecules were significantly elevated in tear fluid of diabetic patients

Statistical *t*-tests further uncovered several endogenous metabolites displaying significantly higher levels in diabetic patients compared to healthy controls (Figure 4). Among them were the essential and non-essential amino acids aspartic acid (Asp), glutamate (Glu), glutamine (Gln), methionine (Met), methionine sulfoxide (MetO), serine (Ser), threonine (Thr), tyrosine (Tyr) and valine (Val) (Figure 4A). Previous metabolomics investigations have already linked these amino acids to insulin resistance and used them as predictors for T2DM risk.[47, 50-52] Moreover, citrulline, ornithine and uric acid were found at significantly higher levels in diabetic patients (Figure 4B), indicating an impaired metabolism (urea cycle and purine catabolism). High levels of uric acid have already been associated with diabetes, however, it is not yet clear whether uric acid contributes to the development of diabetes or if hyperuricemia is a result of insulin resistance.[53] An apparent accumulation of taurine in tears of T2DM patients, as presently observed, has already been described in case of dry eye syndrome.[25] Moreover, taurine levels may be increased in diabetic patients following the long-lasting stress due to the metabolic disorder. Hence, taurine may represent a tear-specific biomarker for T2DM and/ or diabetic retinopathy.

**Figure 4.**
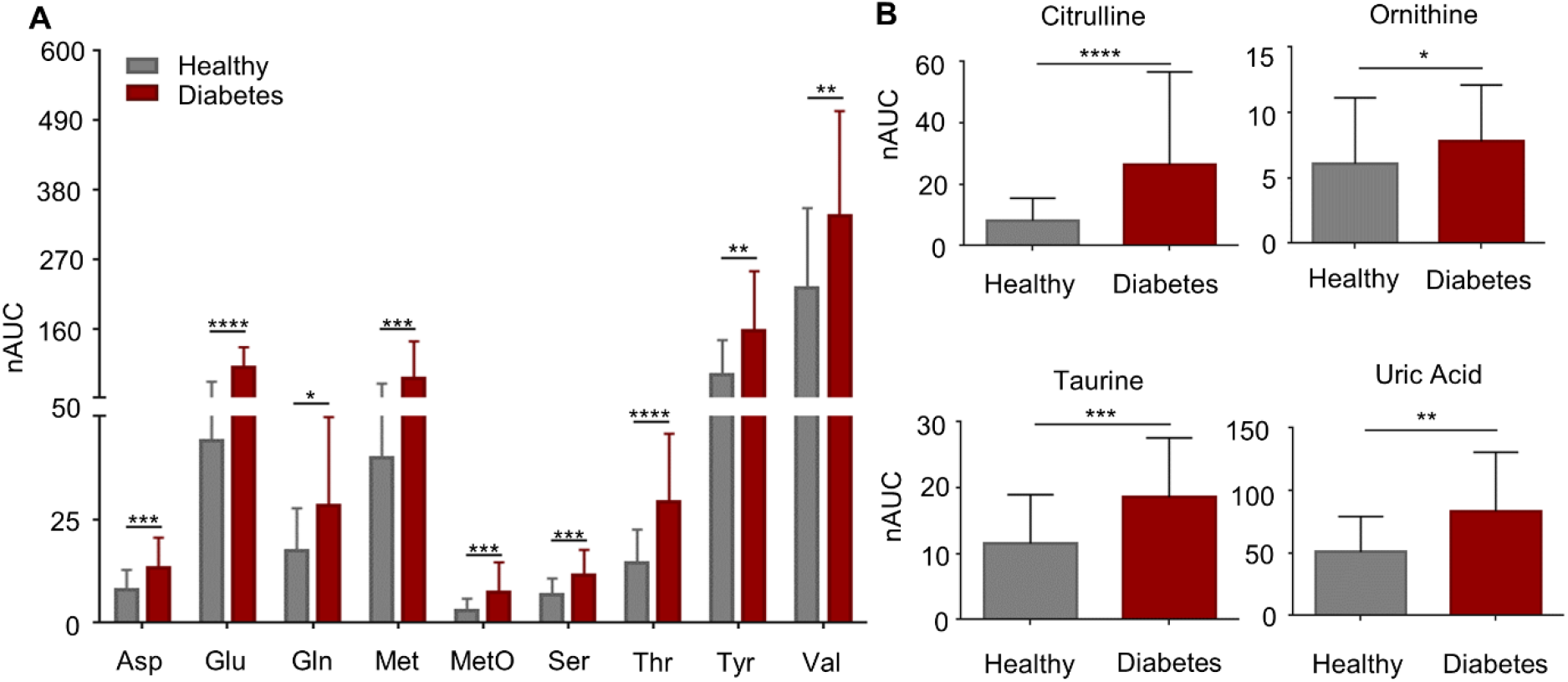
Tear fluid analyses reveals differences in endogenous metabolites that have already been linked to diabetes. **A-B** Box plots of essential and non-essential amino acids that were significantly up-regulated in diabetic patients (n = 16) in contrast to healthy controls (n = 127). Statistical analyses further revealed a significant elevation of citrulline, ornithine, taurine, and uric acid. Normality of the data was checked, but data did not stem from a Gaussian distribution. Two-tailed, unpaired *t*-tests (Mann Whitney Tests) were performed to confirm significance. Boxes represent means and standard deviation. **** = *p*-value < 0.0001, *** = *p*-value < 0.001, ** = *p* -value < 0.01 and * *p*-value < 0.05; Asp, aspartate; Glu, glutamate; Gln, glutamine; Met, methionine, MetO, methionine sulfoxide; nAUC, normalised area under the curve; Ser, serine; Thr, threonine, Tyr, tyrosine; Val, valine.

Conclusively, tear fluid analyses revealed endogenous and exogenous metabolites that were differently regulated or specifically found in diabetic patients. This demonstrates the great potential of LC-MS-based tear fluid analyses for the characterisation of disease state(s) and to support biomarker discovery. This new approach may extend current means to asses individual health states, to identify potential new targets for therapeutic interventions, and to monitor therapeutic efficacy and to control patients’ compliance, which are all factors urgently needed for PPPM.

## Discussion

The present work clearly demonstrates that tear fluid sampled with Schirmer strips can be used for individual metabolic phenotyping. Tears are easily accessible and their collection is non-invasive, painless and fast, thus providing important characteristics regarding point-of-care testing. Compared to blood, the non-invasive nature of tear fluid collection supports repeated sampling of the same individual in order to evaluate dynamic biomarkers which is of great relevance for meaningful metabolic phenotyping. Moreover, a straightforward and quick (<5 min) tear sampling method combined with a fast LC-MS analysis strategy (approximately 7.5 min for metabolites and 20 min for eicosanoids) (Figure 1) offers the opportunity to use tear fluid for large scale longitudinal metabolic studies.

### Distinct metabolic composition of sweat and tear fluid

Since sweat has been already demonstrated to be a rich source for biologically highly relevant molecules[10, 11], this study was conducted to evaluate the composition of tear fluid collected with Schirmer strips in greater detail. Therefore, sweat and tears from 20 healthy participants were collected in parallel in order to investigate and compare the distinct molecular composition of these two non-invasively sampled biofluids. While both biofluids are easily accessible, they also have specific properties: Sweat can be sampled in a highly frequent manner allowing kinetic time-course measurements. Tears are less likely to be contaminated due to the protection of the eyelids and the lipid layer, and the sample volume can be determined, which is necessary for absolute quantification. Numerous endogenous and exogenous small molecule metabolites, which have previously been identified in sweat[10, 11], were also successfully profiled in the minute amounts of tears using the same LC/MS-MS methodology. Interestingly, while some molecules were specifically detected in one biofluid, other molecules showed high abundance differences between the two specimens. For example, kahweol and the whole panel of eicosanoids and eicosanoid-like features were only detected in tear fluid. Molecules such as taurine and epinephrine were found at significantly higher levels in tears compared to sweat (Figure 1). These findings highlight the potential of tear fluid analysis in order to successfully identify low-abundant metabolites, which may be challenging to be detected in other biofluids such as sweat or blood due to matrix effects, pH or polarity. Intriguingly, eicosanoids, important inflammatory mediators were detectable solely in tears demonstrating that tear fluid may be a relevant biofluid allowing insights into disease-related metabolic alterations. Certain diseases often have multiple causes; thus, they may become detectable only at advanced stages due to diagnostic challenges. Earlier molecular diagnosis typically results in better treatment outcome and improved recovery.[54] Most importantly, the more data we can collect on these influencing factors *in vivo* with the help of biofluid analysis, the better we may understand different pathologies. Thus, combining the analysis of several biofluids such as sweat and tears may be valuable to gain complementary information on individual metabolic traits, help to assess individual lifestyle parameters and health states as well as to facilitate a better understanding for different disease pathologies.

### The need for non-invasive metabolic biomonitoring in type 2 diabetes

T2DM is a metabolic disorder that can cause long-term tissue damage and dysfunction of several organs, including the eyes, eventually resulting in sight-loss.[55]. The underlying disease mechanism of T2DM are still not fully elucidated and currently there exists no curative therapy. However, the treatment of pre-diabetes, meaning already impaired β-cell function and insulin resistance but not yet sufficiently high blood sugar levels for diagnosis, has been demonstrated to prevent disease progression.[56] Thus, identifying biomarkers to diagnose the disease prior to clinical manifestation or assess the risk for future insulin resistance and T2DM is crucial in order to initiate preventive treatments by anti-diabetic drugs such as metformin through lifestyle intervention and/ or in time. As already known, ocular diseases (e.g. dry eye disease) as well as systemic diseases (e.g. diabetes) can lead to the accumulation of disease-associated molecules such as proteins or lipids in tear fluid. The fact that T2DM finally results in the development of diabetic retinopathy in 50% of patients[57], justifies tears as a promising source for biological relevant molecules potentially serving as future biomarkers in personalised medicine. So far, most clinical metabolomics studies focusing on the identification of small molecule key players in the pathology of diabetes rely on the analysis of blood/plasma. As metabolite profiles change constantly due to varying environmental demands and disease progression, metabolomic measurements call for repeated analyses in a short timeframe in order to identify dynamic biomarkers. Thus, blood sampling is hardly applicable for time-course analysis due to several compliance issues. Moreover, the lack of inexpensive sampling methods and point-of-care testing devices as well as the absence of high-throughput analysis impede the development of clinical tools used for early disease detection.[54] Non-invasive methods such as tear fluid sampling and analysis seem rather preferable. Since only small volumes of tear fluid can be collected and analytes are generally very low abundant, there is great demand for ultra-sensitive analytical methodologies. With the continuous improvement of detection sensitivity in mass spectrometric approaches, the analysis of the minute amounts of tear fluid and other alternatively sampled biofluids become feasible. Hence, an ultra-sensitive LC-MS method was used for the conduction of a clinical pilot study based on T2DM patients and healthy controls in order to evaluate tears analyses as a diagnostic biofluid for biomarker discovery in T2DM.

### Tear fluid proves as rich source for biomarker discovery in type 2 diabetes mellitus

The opportunity of analysing tear fluid in order to identify potential biomarkers supporting the clinical management of T2DM was presently investigated in a pilot study based on 20 healthy volunteers and 8 T2DM patients. Therefore, the molecular composition of tear fluid was assessed by applying comprehensive LC-MS-based metabolic profiling and subsequently evaluated regarding significant differences between the study cohorts. Altogether, our methodology allowed the identification of 226 metabolites as well as 70 eicosanoids and eicosanoid-like features in tears of all study participants (Figure 2). Statistical analysis between diabetic patients and healthy controls revealed significant difference of certain metabolites but no significant changes in the eicosanoid composition was observed. Regarding metabolites, we were able to detect medications such as metformin, bisoprolol and gabapentin only in tears of diabetic patients as a result of their individual clinical treatment (Figure 3). These findings proofed our analysis strategy as valid approach for further investigation of biomarker discovery. Statistical analyses revealed a significant up-regulation of many amino acids (Asp, Glu, Gln, Met, MetO, Ser, Thr, Tyr, Val) as well as the endogenous metabolites carnitine, citrulline, ornithine, and uric acid in tears of patients suffering from T2DM (Figure 3 and 4). All of these metabolites have already been linked to diabetes and/ or diabetic retinopathy, demonstrating that our results agree with data described in the literature.[40, 47, 48, 52, 58-60] Nonetheless, we have further identified nicotinic acid and taurine to be significantly accumulated in tears of diabetic patients, both have not been reported before and may represent novel biomarker candidates for T2DM, which have to be further evaluated in larger study cohorts.

### The role of branched chain and aromatic amino acids, uric acid and taurine in T2DM

High serum levels of branched chain amino acids (Val, leucine, isoleucine) together with the aromatic amino acids phenylalanine and tyrosine can be used as predictors for identifying individuals at high risk of developing future insulin resistance and T2DM. Levels of these amino acids may be relevant up to 12 years before disease onset.[39, 52] It is speculated that these amino acids act *via* the same pathways as insulin, namely the activation of the mammalian target of rapamycin (mTOR) and its downstream targets. On the other hand, accumulation of these amino acids may cause mitochondrial dysfunction *via* stress kinase stimulation resulting in β-cell apoptosis. Both effects may lead to insulin resistance and T2DM.[61] While elevated serum levels of the respective amino acids have been already reported, we were able to demonstrate the accumulation of many amino acids, i.e. Asp, Glu, Gln, Met, MetO, Ser, Thr, Tyr, Val, also in the tear fluid of patients suffering from T2DM. Similar to amino acids, high serum levels of uric acid were already linked to diabetes but diabetes-related accumulation in tears was not yet reported. Serum hyperuricemia has been strongly associated with pre-diabetes and T2DM since the 1920s, however, it is not clear if accumulation of uric acid is due to reduced kidney function associated with diabetes or if hyperuricemia contributes to diabetic pathology. Yet, it has been shown that uric acid can induce mitochondrial oxidative stress resulting in impaired fatty acid oxidation, insulin-dependent nitric oxide release and glucose delivery.[62] Taurine, which has several important functions including membrane stabilisation, antioxidation properties, osmoregulation and being a pro-inflammatory regulator, was shown to be decreased in plasma levels of diabetic patients in contrast to healthy controls.[25, 63] Here, we have observed contrary results: while decreased plasma taurine levels have been reported, taurine levels in tears were significantly increased in diabetic patients compared to healthy participants. Taurine concentrations were found to be elevated in stressed states, including the inflammation of the ocular surface in dry eye disease.[64] T2DM has also been associated with ocular surface changes and dry eye symptoms, hence, taurine could be a novel biomarker specifically found in tears of diabetic patients reflecting an imbalance of metabolic homeostasis due to long-lasting stress and inflammation of the ocular surface.[65]

### The potential of tear fluid analyses to support PPPM concepts

Metabolic phenotyping of tear fluid derived from TDM patients and healthy controls enabled the successful identification of many meaningful marker molecules potentially serving as future biomarkers. While many of them were already linked to the pathology of diabetes, others seem to be a result of the general metabolic imbalance associated with diabetes. This study clearly demonstrates that tear fluid analyses may not only be used to assess personal health states and lifestyles by evaluating endogenous metabolites but also has the potential to monitor the intake of xenobiotic medications used in the treatment for T2DM in an individualised fashion. Follow-up clinical studies using tear fluid of larger study cohorts are needed in order to evaluate the power of suggested biomarker candidates. Conclusively, tear fluid analyses seems a powerful tool to facilitate the development of novel PPPM-related applications for detecting individuals at risk, monitoring disease progression, therapies and therapeutic efficacy, finding potential targets for therapeutic interventions in conditions such as diabetes, examining lifestyle changes, as well as to check the intake of prescribed medications in clinical practice, all in a non-invasive and painless manner.

## Strength and limitations

Schirmer strip collection was chosen for tear sampling as it is fast, non-invasive, easy, gentle with minimal discomfort for patients, thus, ensuring patients’ compliance and allowing to collect multiple samples from the same participant. Sample preparation of Schirmer strips is straightforward and inexpensive. Nevertheless, Schirmer strip collection could slightly irritate the eye and induce reflex tear secretion in some subjects, and may alter tear fluid composition. Moreover, evaporation of water collected with the strip cannot be excluded, which may result in higher apparent concentrations and may impede the determination of actual collected tear volume and thus, also absolute quantification.[12] We have already demonstrated that the kinetic profiles combined with mathematical models of biochemically related pairs such as caffeine and its metabolites can overcome this normalisation problem and enable the determination of individual sweat flux rates, which can easily be applied to tear fluid analyses as well.[11] Metabolomic profiling of tear fluid using LC-MS/MS is quick, highly sensitive and revealed that tears are a rich source for many endogenous and exogenous metabolites. The presented approach further allowed to identify significant differences between healthy controls and diabetic patients, thereby, demonstrating the applicability of tear fluid analyses as a diagnostic and prognostic tool for clinical applications. However, the sample cohort of this pilot study was quite small, the T2DM sample size was limited, and groups were not age/ BMI matched, thus, other confounders causing these characteristics cannot be fully excluded. Hence, the findings of this study will have to be tested in larger cohorts, which may even allow the identification of further biomarkers and potential targets for therapeutic intervention for diabetes.

## Conclusions and Outlook

We conclude that tear fluid analysis has the great potential to support further developments of PPPM strategies as tears can be sampled non-invasively, gently, repeatedly and thus it allows point-of-care testing. Tears represent a rich source of different classes of endogenous and exogenous metabolites including eicosanoids. Profiling of these metabolites may enable the assessment of individual health states as well as the identification of individual responses to environmental changes and disease states. We have demonstrated that the tear fluid composition of diabetic patients and controls displayed significant differences and presented a panel of potential tear specific biomarkers. Ongoing research may be able to relate dynamic molecular patterns obtained by tear fluid analysis with other disease states or identify individuals at high risk for certain diseases. Hence, we suggest that tear fluid analysis offers many opportunities for applications in precision medicine ranging from biomarker discovery to the monitoring of patients’ compliance and therapeutic efficacy.

## Data Availability

Raw data is secured and available upon reasonable request to the authors

## Acknowledgments

We acknowledge support by the Mass Spectrometry Centre of the Faculty of Chemistry, University of Vienna, and the Joint Metabolome Facility, University of Vienna and Medical University of Vienna. Both facilities are members of the Vienna Life Science Instruments (VLSI). We would also like to thank the Hochschuljubiläumsstiftung (HJS) for their financial support during the course of this research project. The authors are grateful to Azra Smajis for help with the laboratory work. Figure 1A was created with BioRender (www.BioRender.com).

## Additional Information

Supplementary Information

Supplementary Data

### Abbreviations

AA: arachidonic acid
Asp: aspartate
BMI: body mass index
DHA: docosahexaenoic acid
EPA: eicosapentaenoic acid
Gln: glutamine
Glu: glutamate
HbA1c: glycated haemoglobin
HETE: hydroxyeicosatetraenoic acid
LC: liquid chromatography
m/z: mass-to-charge ratio
Met: methionine
MetO: methionine sulfoxide
MS: mass spectrometry
nAUC: normalised area under the curve
PCA: principal component analysis
PPPM: predictive preventive and personalised medicine
ROS: reactive oxygen species
RT: retention time
Ser: serine
T2DM: type 2 diabetes mellitus
Thr: threonine
TNFα: tumour necrosis factor alpha
Tyr: tyrosine
Val: valine

## Declarations

### Availability of data and material

Raw data is secured and available on request.

### Code availability

Not applicable.

### Author Contributions

J.B. performed research, interpreted data, analysed data and wrote the manuscript, A.B. interpreted data and wrote the manuscript, D.S. recruited volunteers and determined clinical parameters, G.H. analysed data, S.M.M. interpreted data and wrote the manuscript, N.H., performed research, A.S., performed research, C.G. conceptualized the project, interpreted data and wrote the manuscript, G.G. conceptualized the project, interpreted data and wrote the manuscript

### Ethics approval

This study was approved by the ethical committee of the Medical University Vienna.

### Consent to participate

Volunteers have given their written, informed consent to participate in this study.

### Consent for publication

Volunteers have given their written, informed consent for publishing the data.

## References

1. Raju KSR, Taneja I, Singh SP, Wahajuddin. Utility of noninvasive biomatrices in pharmacokinetic studies. Biomed Chromatogr. 2013;27(10):1354–66.

2. Li YF, Bouza M, Wu CS, Guo HY, Huang DN, Doron G, et al. Sub-nanoliter metabolomics via mass spectrometry to characterize volume-limited samples. Nat Com. 2020; http://doi.org/10.1038/s41467-020-19444-y.

3. Lesterhuis WJ, Bosco A, Millward MJ, Small M, Nowak AK, Lake RA. Dynamic versus static biomarkers in cancer immune checkpoint blockade: unravelling complexity. Nat Rev Drug Discov. 2017;16(4):264–72.

4. Dutkiewicz EP, Urban PL. Quantitative mass spectrometry of unconventional human biological matrices. Philos Trans Royal Soc A. 2016; http://doi.org/10.1098/rsta.2015.0380.

5. Cheng TT, Zhan XQ. Pattern recognition for predictive, preventive, and personalized medicine in cancer. Epma J. 2017;8(1):51–60.

6. Delgado-Povedano MM, Calderon-Santiago M, Priego-Capote F, de Castro MDL. Development of a method for enhancing metabolomics coverage of human sweat by gas chromatography-mass spectrometry in high resolution mode. Anal Chim Acta. 2016;905:115–25.

7. Zhou L, Beuerman RW. Tear analysis in ocular surface diseases. Prog Retin Eye Res. 2012;31(6):527–50.

8. Ghosh A, Nishtala K. Biofluid lipidome: a source for potential diagnostic biomarkers. Clin Transl Med. 2017; http://doi.org/10.1186/s40169-017-0152-7.

9. Ponzini E, Ami D, Duse A, Santambrogio C, De Palma A, Di Silvestre D, et al. Single-Tear Proteomics: A Feasible Approach to Precision Medicine. Int J Mol Sci. 2021; http://doi.org/10.3390/ijms221910750.

10. Brunmair J, Bileck A, Stimpfl T, Raible F, Del Favero G, Meier-Menches SM, et al. Metabo-tip: a metabolomics platform for lifestyle monitoring supporting the development of novel strategies in predictive, preventive and personalised medicine. Epma J. 2021;12(2):141–53.

11. Brunmair J, Gotsmy M, Niederstaetter L, Neuditschko B, Bileck A, Slany A, et al. Finger sweat analysis enables short interval metabolic biomonitoring in humans. Nat Com. 2021; http://doi.org/10.1038/s41467-021-26245-4.

12. Posa A, Brauer L, Schicht M, Garreis F, Beileke S, Paulsen F. Schirmer strip vs. capillary tube method: Non-invasive methods of obtaining proteins from tear fluid. Ann Anat. 2013;195(2):137–42.

13. Yazdani M, Elgstoen KBP, Rootwelt H, Shahdadfar A, Utheim OA, Utheim TP. Tear Metabolomics in Dry Eye Disease: A Review. Int J Mol Sci. 2019; http://doi.org/10.3390/ijms20153755.

14. Zhou L, Beuerman RW. The power of tears: how tear proteomics research could revolutionize the clinic. Expert Rev Proteomic. 2017;14(3):189–91.

15. Choy CKM, Cho P, Chung WY, Benzie IFF. Water-soluble antioxidants, in human tears: Effect of the collection method. Invest Ophth Vis Sci. 2001;42(13):3130–4.

16. Dartt DA. Tear Lipocalin: Structure and Function. Ocul Surf. 2011;9(3):126–38.

17. Wiesner J, Vilcinskas A. Antimicrobial peptides The ancient arm of the human immune system. Virulence. 2010;1(5):440–64.

18. Li N, Wang N, Zheng J, Liu XM, Lever OW, Erickson PM, et al. Characterization of human tear proteome using multiple proteomic analysis techniques. J Proteome Res. 2005;4(6):2052–61.

19. Rentka A, Koroskenyi K, Harsfalvi J, Szekanecz Z, Szucs G, Szodoray P, et al. Evaluation of commonly used tear sampling methods and their relevance in subsequent biochemical analysis. Ann Clin Biochem. 2017;54(5):521–9.

20. Wu JD, Sigler A, Pfaff A, Cen N, Ercal N, Shi HL. Development of a HPLC-MS/MS method for assessment of thiol redox status in human tear fluids. Anal Biochem. 2021; http://doi.org/10.1016/j.ab.2021.114295.

21. Chen LY, Zhou L, Chan ECY, Neo J, Beuerman RW. Characterization of The Human Tear Metabolome by LC-MS/MS. J Proteome Res. 2011;10(10):4876–82.

22. Pieragostino D, D’Alessandro M, di Ioia M, Di Ilio C, Sacchetta P, Del Boccio P. Unraveling the molecular repertoire of tears as a source of biomarkers: Beyond ocular diseases. Proteom Clin Appl. 2015;9(1-2):169–86.

23. Ambaw YA, Chao C, Ji S, Raida M, Torta F, Wenk MR, et al. Tear eicosanoids in healthy people and ocular surface disease. Sci Rep. 2018; http://doi.org/10.1038/s41598-018-29568-3.

24. Nakatsukasa M, Sotozono C, Shimbo K, Ono N, Miyano H, Okano A, et al. Amino Acid Profiles in Human Tear Fluids Analyzed by High-Performance Liquid Chromatography and Electrospray Ionization Tandem Mass Spectrometry. Am J Ophthalmol. 2011;151(5):799–808.

25. ChenZuo L, Murube J, Latorre A, del Rio RM. Different concentrations of amino acids in tears of normal and human dry eyes. Adv Exp Med Biol. 2002;506:617–21.

26. Hagan S, Tomlinson A, Madden L, Clark AM, Oliver K. Analysis of tear fluid proteins: use of multiplex assays in profiling biomarkers of dry eye disease. Epma J. 2014;5(1):A129.

27. Lam SM, Tong L, Reux B, Duan XR, Petznick A, Yong SS, et al. Lipidomic analysis of human tear fluid reveals structure-specific lipid alterations in dry eye syndrome. J Lipid Res. 2014;55(2):299–306.

28. Kallo G, Emri M, Varga Z, Ujhelyi B, Tozser J, Csutak A, et al. Changes in the Chemical Barrier Composition of Tears in Alzheimer’s Disease Reveal Potential Tear Diagnostic Biomarkers. Plos One. 2016; http://doi.org/10.1371/journal.pone.0158000.

29. Kenny A, Jimenez-Mateos EM, Zea-Sevilla MA, Rabano A, Gili-Manzanaro P, Prehn JHM, et al. Proteins and microRNAs are differentially expressed in tear fluid from patients with Alzheimer’s disease. Sci Rep. 2019; http://doi.org/10.1038/s41598-019-51837-y.

30. Huang Z, Du CX, Pan XD. The use of in-strip digestion for fast proteomic analysis on tear fluid from dry eye patients. Plos One. 2018; http://doi.org/10.1371/journal.pone.0200702.

31. Adigal SS, Rizvi A, Rayaroth NV, John RV, Barik A, Bhandari S, et al. Human tear fluid analysis for clinical applications: progress and prospects. Expert Rev Mol Diagn. 2021;21(8):767–87.

32. Nakajima M, Yamato S, Shimada K, Sato S, Kitagawa S, Honda A, et al. Assessment of drug concentrations in tears in therapeutic drug monitoring: I. Determination of valproic acid in tears by gas chromatography/mass spectrometry with EC/NCI mode. Ther Drug Monit. 2000;22(6):716–22.

33. Sebbag L, Showman L, McDowell EM, Perera A, Mochel JP. Impact of Flow Rate, Collection Devices, and Extraction Methods on Tear Concentrations Following Oral Administration of Doxycycline in Dogs and Cats. J Ocul Pharmacol Th. 2018;34(6):452–9.

34. Vanhaeringen NJ. Clinical Biochemistry of Tears. Surv Ophthalmol. 1981;26(2):84–96.

35. Esmaeli B, Ahmadi MA, Rivera E, Valero V, Hutto T, Jackson DM, et al. Docetaxel secretion in tears -Association with lacrimal drainage obstruction. Arch Ophthalmol. 2002;120(9):1180–2.

36. Hirosawa M, Sambe T, Uchida N, Lee XP, Sato K, Kobayashi S. Determination of nonsteroidal anti-inflammatory drugs in human tear and plasma samples using ultra-fast liquid chromatography-tandem mass spectrometry. Jpn J Ophthalmol. 2015;59(5):364–71.

37. Hagan S, Martin E, Enriquez-de-Salamanca A. Tear fluid biomarkers in ocular and systemic disease: potential use for predictive, preventive and personalised medicine. Epma J. 2016; http://doi.org/10.1186/s13167-016-0065-3.

38. Duarte AA, Mohsin S, Golubnitschaja O. Diabetes care in figures: current pitfalls and future scenario. Epma J. 2018;9(2):125–31.

39. Wishart DS. Emerging applications of metabolomics in drug discovery and precision medicine. Nat Rev Drug Discov. 2016;15(7):473–84.

40. Arneth B, Arneth R, Shams M. Metabolomics of Type 1 and Type 2 Diabetes. Int J Mol Sci. 2019; http://doi.org/10.3390/ijms20102467.

41. Khan MAB, Hashim MJ, King JK, Govender RD, Mustafa H, Al Kaabi J. Epidemiology of Type 2 Diabetes -Global Burden of Disease and Forecasted Trends. J Epidemiol Glob Hea. 2020;10(1):107–11.

42. Golubnitschaja O. Advanced Diabetes Care: Three Levels of Prediction, Prevention & Personalized Treatment. Curr Diabetes Rev. 2010;6(1):42–51.

43. Peddinti G, Cobb J, Yengo L, Froguel P, Kravic J, Balkau B, et al. Early metabolic markers identify potential targets for the prevention of type 2 diabetes. Diabetologia. 2017;60(9):1740–50.

44. Niederstaetter L, Neuditschko B, Brunmair J, Janker L, Bileck A, Del Favero G, et al. Eicosanoid Content in Fetal Calf Serum Accounts for Reproducibility Challenges in Cell Culture. Biomolecules. 2021; http://doi.org/10.3390/biom11010113.

45. Neuditschko B, Leibetseder M, Brunmair J, Hagn G, Skos L, Gerner MC, et al. Epithelial Cell Line Derived from Endometriotic Lesion Mimics Macrophage Nervous Mechanism of Pain Generation on Proteome and Metabolome Levels. Biomolecules. 2021; http://doi.org/10.3390/biom11081230.

46. Tyanova S, Temu T, Sinitcyn P, Carlson A, Hein MY, Geiger T, et al. The Perseus computational platform for comprehensive analysis of (prote)omics data. Nat Methods. 2016;13(9):731–40.

47. Strand E, Rebnord EW, Flygel MR, Lysne V, Svingen GFT, Tell GS, et al. Serum Carnitine Metabolites and Incident Type 2 Diabetes Mellitus in Patients With Suspected Stable Angina Pectoris. J Clin Endocr Metab. 2018;103(3):1033–41.

48. Sun L, Liang LM, Gao XF, Zhang HP, Yao P, Hu Y, et al. Early Prediction of Developing Type 2 Diabetes by Plasma Acylcarnitines: A Population-Based Study. Diabetes Care. 2016;39(9):1563–70.

49. Sena CM, Bento CF, Pereira P, Seica R. Diabetes mellitus: new challenges and innovative therapies. Epma J. 2010;1(1):138–63.

50. Alqudah A, Wedyan M, Qnais E, Jawarneh H, McClements L. Plasma Amino Acids Metabolomics’ Important in Glucose Management in Type 2 Diabetes. Front Pharmacol. 2021; http://doi.org/10.3389/fphar.2021.695418.

51. Sun Y, Gao HY, Fan ZY, He Y, Yan YX. Metabolomics Signatures in Type 2 Diabetes: A Systematic Review and Integrative Analysis. J Clin Endocrinol Metab. 2020; http://doi.org/10.1210/clinem/dgz240.

52. Wang TJ, Larson MG, Vasan RS, Cheng S, Rhee EP, McCabe E, et al. Metabolite profiles and the risk of developing diabetes. Nat Med. 2011;17(4):448–53.

53. Dehghan A, van Hoek M, Sijbrands EJ, Hofman A, Witteman JC. High serum uric acid as a novel risk factor for type 2 diabetes. Diabetes Care. 2008;31(2):361–2.

54. Zhang AH, Sun H, Wang XJ. Saliva Metabolomics Opens Door to Biomarker Discovery, Disease Diagnosis, and Treatment. Appl Biochem Biotech. 2012;168(6):1718–27.

55. Lechner J, O’Leary OE, Stitt AW. The pathology associated with diabetic retinopathy. Vision Res. 2017;139:7–14.

56. Khan RMM, Chua ZJY, Tan JC, Yang YY, Liao ZH, Zhao Y. From Pre-Diabetes to Diabetes: Diagnosis, Treatments and Translational Research. Medicina. 2019;http://doi.org/10.3390/medicina55090546.

57. Fong DS, Aiello L, Gardner TW, King GL, Blankenship G, Cavallerano JD, et al. Retinopathy in diabetes. Diabetes Care. 2004;http://doi.org/10.2337/diacare.27.2007.s84.

58. Ahola-Olli AV, Mustelin L, Kalimeri M, Kettunen J, Jokelainen J, Auvinen J, et al. Circulating metabolites and the risk of type 2 diabetes: a prospective study of 11,896 young adults from four Finnish cohorts. Diabetologia. 2019;62(12):2298–309.

59. Sumarriva K, Uppal K, Ma CY, Herren DJ, Wang YT, Chocron IM, et al. Arginine and Carnitine Metabolites Are Altered in Diabetic Retinopathy. Invest Ophth Vis Sci. 2019;60(8):3119–26.

60. Long JL, Yang ZR, Wang L, Han YM, Peng C, Yan C, et al. Metabolite biomarkers of type 2 diabetes mellitus and pre-diabetes: a systematic review and meta-analysis. Bmc Endocr Disord. 2020; http://doi.org/10.1186/s12902-020-00653-x.

61. Yoon MS. The Emerging Role of Branched-Chain Amino Acids in Insulin Resistance and Metabolism. Nutrients. 2016; http://doi.org/10.3390/nu8070405.

62. King C, Lanaspa MA, Jensen T, Tolan DR, Sanchez-Lozada LG, Johnson RJ. Uric Acid as a Cause of the Metabolic Syndrome. Contrib Nephrol. 2018;192:88–102.

63. Sak D, Erdenen F, Muderrisoglu C, Altunoglu E, Sozer V, Gungel H, et al. The Relationship between Plasma Taurine Levels and Diabetic Complications in Patients with Type 2 Diabetes Mellitus. Biomolecules. 2019; http://doi.org/10.3390/biom9030096.

64. ChenZhuo L, Murube J, Latorre A, del Rio RM. Different concentrations of amino acids in tears of normal and human dry eyes. Adv Exp Med Biol. 2002;506:617–21.

65. He FL, Zhao ZL, Liu Y, Lu LN, Fu Y. Assessment of Ocular Surface Damage during the Course of Type 2 Diabetes Mellitus. J Ophthalmol. 2018; http://doi.org/10.1155/2018/1206808.

