## Supplementary Information for "Metabolic phenotyping of tear fluid as a prognostic tool for personalised medicine exemplified by T2DM patients"

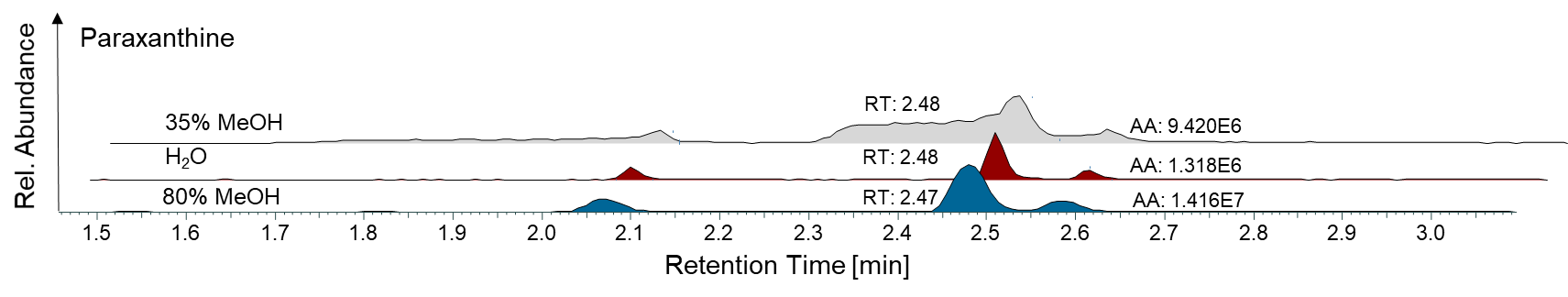

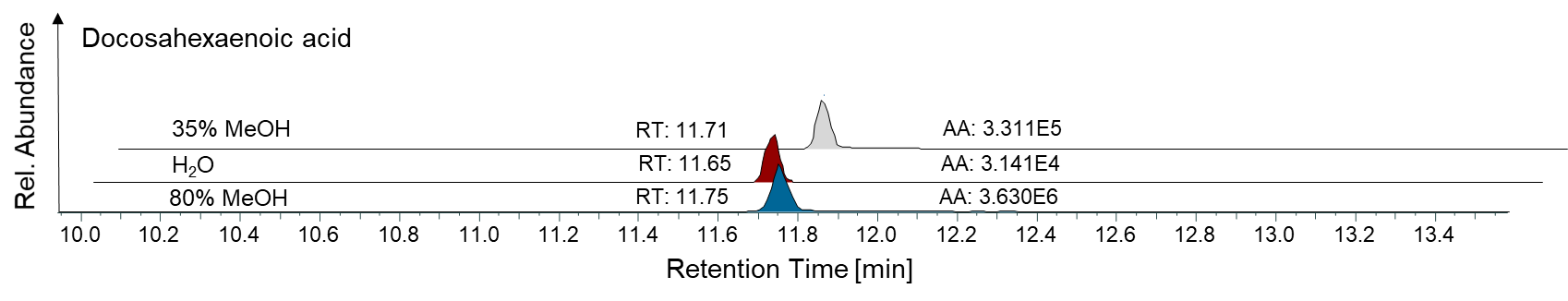

**Supplementary Figure 1**. Respective peak areas of paraxanthine (RT = 2.47 min) and docosahexaenoic acid (RT = 11.75 min) extracted from tear fluid with water, 35% methanol and 80% methanol. AA, absolute area; RT, retention time

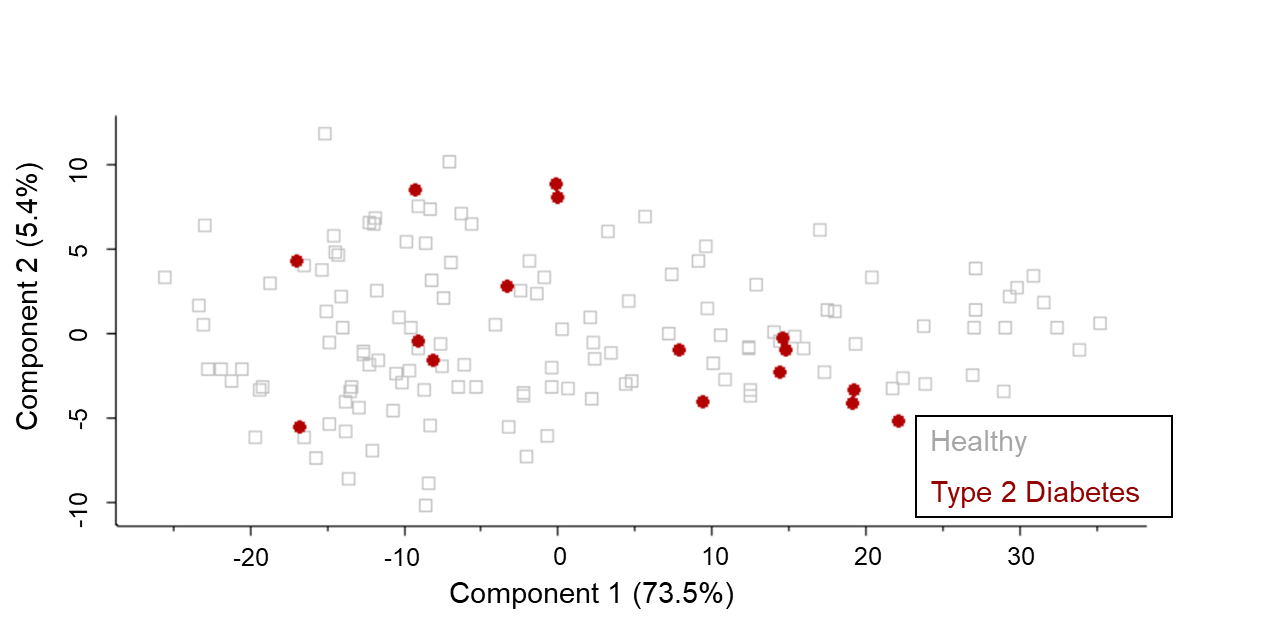

**Supplementary Figure 2.** Principal component analyses calculated with a set of 70 eicosanoids. No clear distinction between healthy controls and diabetic patients was observed.

**Supplementary Table 1:** Internal standards for the standard mixture and their respective concentrations in each sample.

| **Name** | **Abbreviation** | **c [pg/µL]** |
| --- | --- | --- |
| 12S-Hydroxyeicosatetraenoic acid-d8 | 12S-HETE-d8 | 6.67 |
| 15S-Hydroxyeicosatetraenoic acid-d8 | 15S-HETE-d8 | 6.67 |
| 5-Oxo-Eicosatetraenoic acid-d7 | 5-Oxo-ETE-d7 | 20 |
| 11,12-Dihydroxy-5,8,14-eicosatrienoic acid-d11 | 11,12-DiHETrE-d11 | 6.67 |
| Prostaglandin E2-d4 | PGE2-d4 | 13.33 |
| 20-Hydroxyeicosatetraenoic acid-d6 | 20-HETE-d6 | 6.67 |

**Supplementary Table 2:** Inclusion list of 31 eicosanoids and their precursors used for mass spectrometric analysis

| **Mass [m/z]** | **Mass [m/z]** | **Mass [m/z]** | **Mass [m/z]** |
| --- | --- | --- | --- |
| 254.2245 | 301.2168 | 327.2781 | 351.2177 |
| 275.2011 | 303.2330 | 329.2480 | 353.2328 |
| 277.2167 | 311.2228 | 333.2071 | 355.2428 |
| 279.2324 | 315.1966 | 335.2222 | 357.2585 |
| 281.2480 | 317.2122 | 337.2384 | 359.2222 |
| 283.2637 | 319.2279 | 343.2279 | 367.3576 |
| 293.2122 | 321.2435 | 348.3069 | 375.2171 |
| 295.2279 | 327.2324 | 349.2020 |  |
